# Machine learning-based classification of Alzheimer’s disease and its at-risk states using personality traits, anxiety, and depression

**DOI:** 10.1101/2022.11.30.22282930

**Authors:** Konrad F. Waschkies, Joram Soch, Margarita Darna, Anni Richter, Slawek Altenstein, Aline Beyle, Frederic Brosseron, Friederike Buchholz, Michaela Butryn, Laura Dobisch, Michael Ewers, Klaus Fliessbach, Tatjana Gabelin, Wenzel Glanz, Doreen Goerss, Daria Gref, Daniel Janowitz, Ingo Kilimann, Andrea Lohse, Matthias H. Munk, Boris-Stephan Rauchmann, Ayda Rostamzadeh, Nina Roy, Eike Jakob Spruth, Peter Dechent, Michael T. Heneka, Stefan Hetzer, Alfredo Ramirez, Klaus Scheffler, Katharina Buerger, Christoph Laske, Robert Perneczky, Oliver Peters, Josef Priller, Anja Schneider, Annika Spottke, Stefan Teipel, Emrah Düzel, Frank Jessen, Jens Wiltfang, Björn H. Schott, Jasmin M. Kizilirmak

**Affiliations:** German Center for Neurodegenerative Diseases (DZNE), Göttingen, Germany; Department of Psychiatry and Psychotherapy, University Medical Center Göttingen, Göttingen, Germany; Bernstein Center for Computational Neuroscience, Berlin, Germany; Leibniz Institute for Neurobiology, Magdeburg, Germany; German Center for Neurodegenerative Diseases (DZNE), Berlin, Germany; Department of Psychiatry and Psychotherapy, Charité, Berlin, Germany; German Center for Neurodegenerative Diseases (DZNE), Bonn, Germany; University of Bonn Medical Center, Dept. of Neurodegenerative Disease and Geriatric Psychiatry/Psychiatry, Bonn, Germany; Charité – Universitätsmedizin Berlin, corporate member of Freie Universität Berlin and Humboldt-Universität zu Berlin-Institute of Psychiatry and Psychotherapy; German Center for Neurodegenerative Diseases (DZNE), Magdeburg, Germany; German Center for Neurodegenerative Diseases (DZNE), Munich, Germany; Institute for Stroke and Dementia Research (ISD), University Hospital, LMU Munich, Munich, Germany; German Center for Neurodegenerative Diseases (DZNE), Rostock, Germany; Department of Psychosomatic Medicine, Rostock University Medical Center, Rostock, Germany; German Center for Neurodegenerative Diseases (DZNE), Tübingen, Germany; Section for Dementia Research, Hertie Institute for Clinical Brain Research and Department of Psychiatry and Psychotherapy, University of Tübingen, Tübingen, Germany; Department of Psychiatry and Psychotherapy, University Hospital, LMU Munich, Munich, Germany; Sheffield Institute for Translational Neuroscience (SITraN), University of Sheffield, Sheffield, UK; Department of Neuroradiology, University Hospital LMU, Munich, Germany; Department of Psychiatry, University of Cologne, Medical Faculty, Cologne, Germany; MR-Research in Neurosciences, Department of Cognitive Neurology, Georg-August-University Goettingen, Germany; Berlin Center for Advanced Neuroimaging, Charité – Universitätsmedizin Berlin, Berlin, Germany; Excellence Cluster on Cellular Stress Responses in Aging-Associated Diseases (CECAD), University of Cologne, Cologne, Germany; Division of Neurogenetics and Molecular Psychiatry, Department of Psychiatry and Psychotherapy, Faculty of Medicine and University Hospital Cologne, University of Cologne, Cologne, Germany; Department of Psychiatry & Glenn Biggs Institute for Alzheimer’s and Neurodegenerative Diseases, San Antonio, TX, USA; Department for Biomedical Magnetic Resonance, University of Tübingen, 72076 Tübingen, Germany; Munich Cluster for Systems Neurology (SyNergy) Munich, Munich, Germany; Ageing Epidemiology Research Unit (AGE), School of Public Health, Imperial College London, London, UK; School of Medicine, Technical University of Munich; Department of Psychiatry and Psychotherapy, Munich, Germany; University of Edinburgh and UK DRI, Edinburgh, United Kingdom; Department of Neurology, University of Bonn, Bonn, Germany; Institute of Cognitive Neurology and Dementia Research (IKND), Otto-von-Guericke University, Magdeburg, Germany; Neurosciences and Signaling Group, Institute of Biomedicine (iBiMED), Department of Medical Sciences, University of Aveiro, Aveiro, Portugal; Neurodidactics and NeuroLab, Institute for Psychology, University of Hildesheim, Hildesheim, Germany; German Center for Mental Health (DZPG); Center for Intervention and Research on adaptive and maladaptive brain Circuits underlying mental health (C-I-R-C), Jena-Magdeburg-Halle

**Keywords:** Alzheimer’s disease, subjective cognitive decline, amnestic mild cognitive impairment, biomarker, cerebrospinal fluid, personality, fMRI, resting-state, support vector machine, machine learning

## Abstract

**Background:** Alzheimer’s disease (AD) is often preceded by stages of cognitive impairment, namely subjective cognitive decline (SCD) and mild cognitive impairment (MCI). While cerebrospinal fluid (CSF) biomarkers are established predictors of AD, other non-invasive candidate predictors include personality traits, anxiety, and depression, among others. These predictors offer non-invasive assessment and exhibit changes during AD development and preclinical stages.

**Methods:** In a cross-sectional design, we comparatively evaluated the predictive value of personality traits (Big Five), geriatric anxiety and depression scores, resting-state functional magnetic resonance imaging activity of the default mode network, apoliprotein E (ApoE) genotype, and CSF biomarkers (tTau, pTau181, Aβ42/40 ratio) in a multi-class support vector machine classification. Participants included 189 healthy controls (HC), 338 individuals with SCD, 132 with amnestic MCI, and 74 with mild AD from the multicenter DZNE-Longitudinal Cognitive Impairment and Dementia Study (DELCODE).

**Results:** Mean predictive accuracy across all participant groups was highest when utilizing a combination of personality, depression, and anxiety scores. HC were best predicted by a feature set comprised of depression and anxiety scores and participants with AD were best predicted by a feature set containing CSF biomarkers. Classification of participants with SCD or aMCI was near chance level for all assessed feature sets.

**Conclusion:** Our results demonstrate predictive value of personality trait and state scores for AD. Importantly, CSF biomarkers, personality, depression, anxiety, and ApoE genotype show complementary value for classification of AD and its at-risk stages.

**Key Points:**

- Multi-class support vector machine classification was used to compare the predictive value of well-established and non-invasive, easy-to-assess candidate variables for classifying participants with healthy cognition, subjective cognitive decline, amnestic mild cognitive impairment, and mild Alzheimer’s disease.
- Personality traits, geriatric anxiety and depression scores, resting-state functional magnetic resonance imaging activity of the default mode network, ApoE genotype, and CSF biomarkers were comparatively evaluated.
- A combination of personality, anxiety, and depression scores provided the highest predictive accuracy, comparable to CSF biomarkers, indicating complementary value.
- Established and candidate predictors had limited success in classifying SCD and aMCI, underscoring the heterogeneity of these cognitive states and emphasizing the need for standardizing terminology and diagnostic criteria.

## 1 Introduction

Alzheimer’s disease (AD) is commonly preceded by cognitive impairment states, namely subjective cognitive decline (SCD) and mild cognitive impairment (MCI). While MCI requires a measurable deviation from normal cognitive performance as assessed by neuropsychological testing, SCD does not. As both are recognized risk factors for AD (Albert et al. 2011; Jessen et al. 2014), effective treatment for AD requires early intervention (Blennow et al. 2010; Sperling et al. 2011; Binnewijzend et al. 2012; Buchhave et al. 2012; Jessen et al. 2014; Badhwar et al. 2017; Jessen et al. 2018).

Established biomarkers for the diagnosis of AD and associated risk stages are altered levels of amyloid beta (Aβ1-42), total tau (tTau), and phosphorylated tau (pTau181) in cerebrospinal fluid (CSF; Blennow et al. 2010; Olsson et al. 2016; Badhwar et al. 2017). Obtaining CSF samples requires an invasive lumbar puncture and is typically only performed in cases of clinical suspicion. Hence, less invasive measures have been proposed. This study undertook a comparative assessment of the predictive value of voxel-wise resting-state functional magnetic resonance imaging activity of the default mode network (DMN), personality traits, depression, anxiety, apolipoprotein E (ApoE) genotype, and CSF biomarkers. These predictors were employed in a machine-learning classification framework to distinguish between different groups of participants positioned along the trajectory of Alzheimer’s disease or those in a cognitively healthy state (Figure 1).

**Figure 1.**
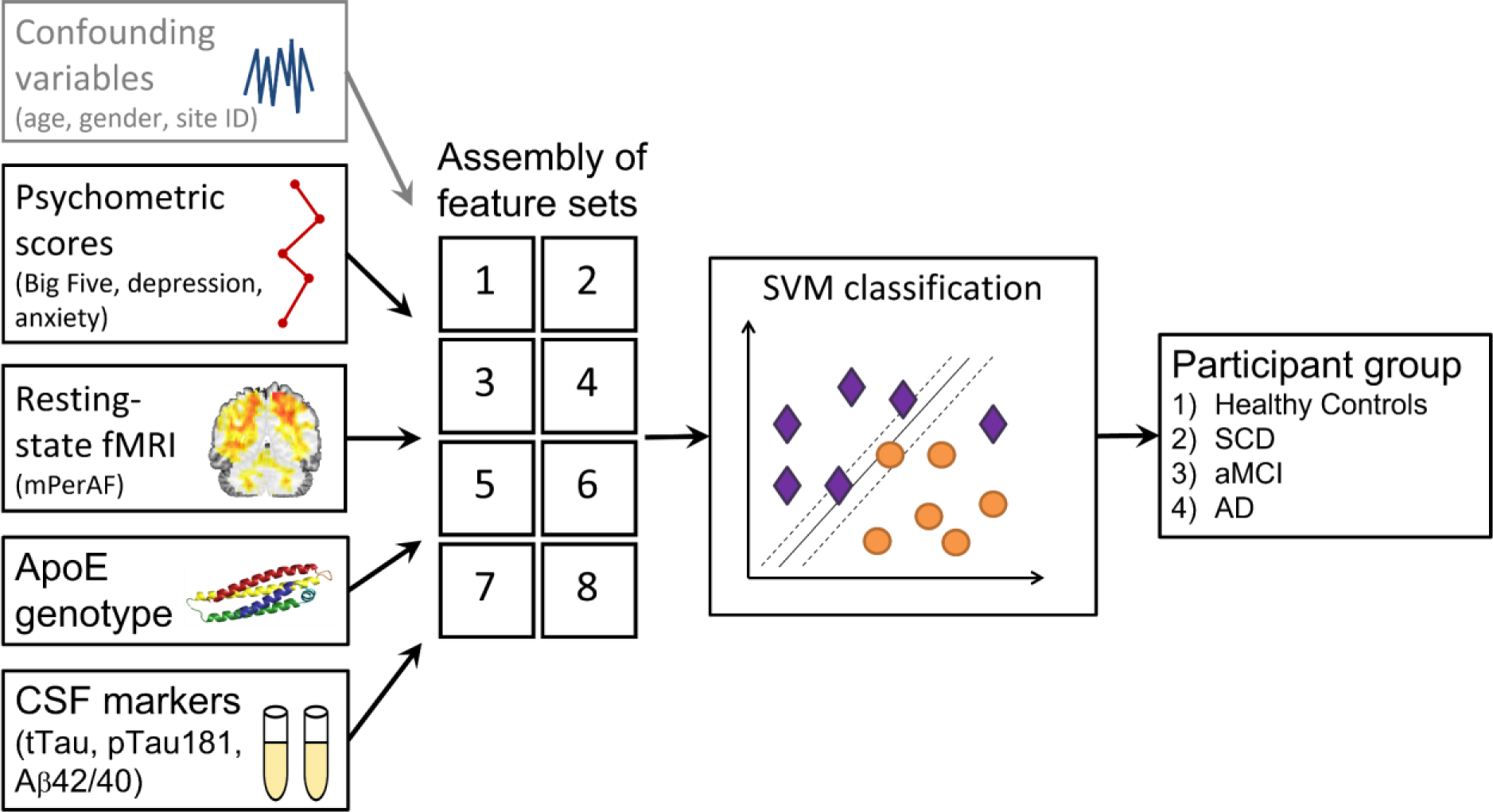
Study design. In a cross-sectional design, predictor variables were combined into feature sets that were used in the SVM classification to predict participant groups. The feature set “confounding variables” was included in all other feature sets and also served as the base model.

At an intra-individual level, personality traits (McCrae & Costa 1987) change in premorbid cognitive states and in AD itself. Overall, neuroticism has been observed to increase during the transition from normal cognition to amnestic MCI, while extraversion, openness, and conscientiousness decrease, with limited evidence for lower agreeableness (Mendez Rubio et al. 2013; Yoneda et al. 2016; Terracciano et al. 2017; Caselli et al. 2018). Similarly, at an inter-individual level, individuals with AD display higher neuroticism and lower scores in agreeableness, extraversion, conscientiousness, and openness compared to healthy controls in both self- and informant ratings (Duchek et al. 2007; Robins Wahlin and Byrne 2011). In general, a linear trend reflecting the severity of cognitive decline is apparent in personality trait scores, indicating that alterations in AD are more notable and pronounced compared to its preceding stages.

Personality traits are considered rather stable throughout life, while anxiety and depression are transient states. However, anxiety and depression are widely reported to correlate with personality traits (Kotov et al. 2010; Klein et al. 2011; Hakulinen et al. 2015) and may be regarded as proxies for neuroticism (Costa Jr. and McCrae 2008; Soto et al. 2009). Higher levels of depression and anxiety are consistently associated with subjective cognitive decline (SCD) (Hill et al. 2016), aMCI (Ismail et al. 2017; Mirza et al. 2017), and AD (Leung et al. 2021) and may be used as predictors for these cognitive states. Comparisons of affective symptoms between SCD/MCI and SCD/AD have yielded inconsistent results, but higher prevalence of depressive symptoms is observed compared to healthy controls (Hill et al. 2016). Higher anxiety and depression levels increase the risk of converting from (a)MCI to AD (Palmer et al. 2007; Li et al. 2016; Li et al. 2018; Peakman et al. 2020) and treatment of these conditions might potentially reduce the conversion rate (Cooper et al. 2015). Additionally, the rate of cognitive decline is reported to be influenced by the age of depression onset (Ly et al. 2021). There is ongoing debate regarding whether depression constitutes a risk factor or an initial manifestation of AD, or both (Panza et al. 2010; Singh-Manoux et al. 2017; Invernizzi et al. 2021).

Activity of the DMN (Raichle et al. 2001) can be assessed employing resting-state fMRI (Andrews-Hanna et al. 2014) and metrics like PerAF (Jia et al. 2020) by measuring BOLD signal fluctuations. Patterns of AD-typical Aβ plaques deposition and disturbances in DMN functional connectivity of the DMN show considerable overlap (Mohan et al. 2016). DMN functional alterations have been described in individuals with aMCI and AD for a range of measures, including amplitude of low frequency fluctuations, therefore holding potential diagnostic value for identifying AD and its at-risk states (Blennow et al. 2010; Mevel et al. 2011; Cha et al. 2013; Badhwar et al. 2017).

The ε4 allele in the apolipoprotein E (ApoE) gene is a genetic risk factor for AD, showing a gene-dose effect of the ApoE ε4 risk allele, with ApoE ε4 homozygotes having a higher risk than ApoE ε3/ε4 heterozygotes (Blennow et al. 2010; Sperling et al. 2011; Dubois et al. 2014; Jansen et al. 2015; Hansson et al. 2018; Jessen et al. 2018; Leuzy et al. 2021). The ApoE genotype is proposed as a risk marker in individuals with SCD (Jessen et al. 2014).

Previous research has mostly tested the aforementioned predictors individually in discriminating cognitively healthy individuals from those at-risk for or with AD. Here, we assessed their diagnostic value in a cross-sectional multi-class classification approach (Ramzan et al. 2019), including all four participant groups simultaneously. Our primary focus was to evaluate the role of personality traits, both individually and in combination with depression and anxiety. Furthermore, we aimed to compare the performance of all assessed feature sets in terms of their respective predictive^1^ accuracies, i.e., class and decoding accuracies.

Our hypotheses were as follows:

1. Measures of personality traits would yield significant predictive accuracies above chance across all participant groups.
2. Combining personality traits, depression, and anxiety scores would improve predictive accuracies compared to personality traits alone.
3. A feature set comprising non-invasive predictors (voxel-wise resting-state activity of the DMN, personality traits, depression and anxiety scores, and ApoE genotype) would yield equal or higher predictive accuracies across all groups compared to a feature set consisting of CSF biomarkers (tTau, pTau181, and Aβ42/40 ratio).

## 2 Materials and Methods

### 2.1 Participants

For our cross-sectional study, we used baseline data from participants recruited through the DELCODE study. For detailed information on the DELCODE study, see Jessen et al. (2018). We included a large cohort of 733 participants that were assigned to four different groups based on their entry diagnosis: HC, SCD, aMCI, and mild AD. All participants were aged 60 years or older, fluent in German, able to give informed consent, and had a study partner present.

Participants for the study were recruited either through local newspaper advertisements or from memory clinics. Healthy controls self-identified as cognitively healthy and passed a telephone screening for SCD. These individuals were included as HC if their memory test performance was within 1.5 standard deviations (SD) of the age-, gender-, and education-adjusted normal performance on all Consortium to Establish a Registry for Alzheimer’s Disease (CERAD) subtests and if they did not meet the SCD criteria (Jessen et al. 2014). Conversely, individuals expressing cognitive decline concerns to the memory center physician were categorized as either SCD or aMCI, based on a comprehensive semi-structured interview following the SCD-plus criteria (Jessen et al. 2014) and their CERAD performance. SCD participants outperformed the -1.5 SD below normal, while aMCI patients underperformed (>1.5 SD) on the “recall word list” subtest, thus excluding non-amnestic MCI participants. They did not meet the criteria for dementia, and their inclusion was based on the memory clinic diagnoses, which adhered to the current research criteria for MCI as defined by the National Institute on Aging-Alzheimer’s Association (Albert et al. 2011; McKhann et al. 2011).

Assignment to the AD group was based on both clinical diagnosis and on the Mini Mental Status Examination (MMSE). Only participants with mild AD (>18 points and <26 points on the MMSE) were included. Aside from HC, all participant groups (SCD, MCI, AD) were memory clinic referrals and underwent clinical assessments at their respective memory centers. These assessments consisted of a medical history review, psychiatric and neurological examinations, neuropsychological testing, blood laboratory analysis, and routine MRI scans. Cognitive function was measured using the CERAD neuropsychological test battery, which was administered at all memory centers.

### 2.2 MRI data acquisition

Structural and functional MRI data were acquired on 3T Siemens scanners following the DELCODE study protocol (Jessen et al. 2018; Düzel et al. 2019). A T1-weighted MPRAGE image (TR = 2.5 s, TE = 4.37 ms, flip-α = 7°; 192 slices, 256 x 256 in-plane resolution, voxel size = 1 x 1 x 1 mm) was acquired for co-registration and improved spatial normalization.

The MPRAGE was followed by a 7:54 min resting-state fMRI (rs-fMRI) acquisition, during which T2*-weighted echo-planar images (EPI; TR = 2.58 s, TE = 30 ms, flip-α = 80°; 47 axial slices, 64 x 64 in-plane resolution, voxel size = 3.5 x 3.5 x 3.5 mm) were acquired in odd-even interleaved-ascending slice order. Participants were instructed to lie inside the scanner with eyes closed, but without falling asleep. Directly after, phase and magnitude fieldmap images were acquired to improve correction for artifacts resulting from magnetic field inhomogeneities via unwarping. This was followed by brief co-planar T1-weighted inversion recovery EPIs. The complete study protocol included other scanning sequences not used in the analyses reported here (Jessen et al. 2018).

### 2.3 fMRI data preprocessing and analysis

Data preprocessing and computation of mPerAF maps were performed using Statistical Parametric Mapping (SPM12; Wellcome Trust Center for Neuroimaging, University College London, London, UK) and the RESTplus toolbox (Jia et al. 2019), following a recently described protocol (Kizilirmak et al. 2022). EPIs were corrected for acquisition time delay (*slice timing*), head motion (*realignment*), and magnetic field inhomogeneities (*unwarping*), using voxel-displacement maps (VDMs) derived from the fieldmaps. The MPRAGE image was spatially co-registered to the mean unwarped image and *segmented* into six tissue types, using the unified segmentation and normalization algorithm implemented in SPM12. The resulting forward deformation parameters were used to *normalize unwarped* EPIs into a standard stereotactic reference frame (Montreal Neurological Institute, MNI; voxel size = 3 x 3 x 3 mm). Normalized images were spatially *smoothed* using an isotropic Gaussian kernel of 6 mm full width at half maximum.

PerAF is a voxel-wise, scale-independent measure of low-frequency (0.01-0.08 Hz) BOLD signal fluctuations relative to the mean BOLD signal intensity for each time point, averaged across the whole time series (Jia et al. 2020). The global-mean-adjusted PerAF (mPerAF) was computed from rs-fMRI using an adapted version^2^ of the *RESTplus* toolbox (Jia et al. 2019). A DMN mask (Shirer et al. 2012) was applied, representing a composite of functionally defined regions of interest (ROIs), and the resulting mPerAF maps served as voxel-wise mean-centered predictor variables.

### 2.4 Clinical and risk factor assessments

Trained study physicians administered the baseline clinical assessments in the DELCODE study. These assessments followed a fixed order and were completed within a single day. Caregivers of participants with AD were allowed to help complete the questionnaires. Clinical assessments included the Geriatric Depression Scale short form (GDS; Sheikh & Yesavage 1986), the Geriatric Anxiety Inventory short form (GAI-SF; Byrne & Pachana 2011), and the Big Five Inventory short form (BFI-10; Rammstedt & John 2007; Rammstedt et al. 2017). Scores on the five personality scales (each calculated as the mean of the two respective items) were included as five standardized predictors. The sum scores of GDS and GAI-SF were included as standardized predictors, respectively.

### 2.5 ApoE genotyping

The single nucleotide polymorphisms (SNPs) rs7412 and rs429358, which define the ε2, ε3, and ε4 alleles of the ApoE gene, were determined using a TaqMan® SNP Genotyping Assay (ThermoFisher Scientific). ApoE ε4 non-carriers (ε2/ε2, ε2/ε3, ε3/ε3) were coded as 0, heterozygotes (ε2/ε4, ε3/ε4) were coded as 1, and homozygotes (ε4/ε4) were coded as 2.

### 2.6 Cerebrospinal fluid biomarker assessment

Cerebrospinal fluid biomarkers (tTau, pTau181, and Aβ42/40 ratio; collectively referred to as CSF biomarkers) were measured using commercially available kits according to manufacturers’ specifications: V-PLEX Aβ Peptide Panel 1 (6E10) Kit (K15200E) and V-PLEX Human Total Tau Kit (K151LAE) (Mesoscale Diagnostics LLC, Rockville, USA), and Innotest Phospho-Tau(181P) (81581; Fujirebio Germany GmbH, Hannover, Germany).

### 2.7 Assessment of confounding features

Chronological age was included as a standardized predictor (mean = 0, SD = 1). The acquisition site predictor used in the DELCODE study included ten distinct sites across Germany, which were represented as dummy-coded predictors using ten binary variables. Gender was included as a dummy-coded predictor with two binary predictors.

### 2.8 Prediction of outcome from predictor variables and performance assessment

Predictor variables were combined into eight feature sets (Figure 1). In this study, we will employ the terms “predictor(s)” and “feature(s)” interchangeably, as well as “group(s)” and “class(es)”, to represent the same concept.

1. **Base model**: age, gender, site
2. **mPerAF**: base model, mPerAF
3. **Personality**: base model, BFI-10
4. **Depression, anxiety**: base model, GDS, GAI-SF
5. **Personality extended**: base model, BFI-10, GDS, GAI-SF
6. **ApoE**: base model, ApoE genotype
7. **CSF**: base model, tTau, pTau181, Aβ42/40 ratio
8. **All w/o CSF**: base model, mPerAF, BFI-10, GDS, GAI-SF, ApoE genotype

To predict the outcome variable (participant group) with the feature sets, we employed Support Vector Classification (SVC) using linear Support Vector Machines (SVMs) with soft-margin parameter *C* = 1 and 10-fold cross-validation. All SVM analyses were implemented using LibSVM in MATLAB via custom scripts available on GitHub (https://github.com/JoramSoch/ML4ML).

Predictive performance of participant classification was assessed using decoding accuracy (DA), that is, the average proportion of correctly classified participants across all groups, and class accuracy (CA), that is, the same proportion, separately for each group, each ranging between 0 and 1.

For each feature set, statistically significant differences from chance-level prediction for DA and CAs were tested, and pairwise comparisons of each feature set against the base model were performed. This was done using one-tailed paired *t*-tests for the classification performance of each feature set against the base model, with each pair consisting of a subsample evaluated using both feature sets. Bonferroni-Holm correction was applied for multiple testing. Additionally, a subsample-by-subsample correlation matrix of DAs across all permutations was computed and incorporated into a general linear model of the pairwise accuracy differences across all subsamples. All scripts used to perform the analyses are available at https://github.com/jmkizilirmak/DELCODE162.

### 2.9 Handling of missing values and unbalanced class sizes

Participants with missing data for age, gender, site, mPerAF, BFI-10, GDS, GAI-SF, and ApoE genotype were excluded from analysis (N = 663; 179 HC, 308 SCD, 113 aMCI, 63 AD). Due to additional missing CSF biomarker values, additional exclusions were made for the “CSF” feature set (N = 341; 75 HC, 155 SCD, 71 aMCI, 40 AD) and the “CSF” feature set was excluded from inferential comparisons to maintain statistical power. Supplementary information provides an alternative analysis with equal sample sizes (N = 311; Table S4) across all feature sets, as well as an analysis with SCD and aMCI groups merged into an “at-risk for AD” group (Table S2).

Subsampling was used to ensure equal numbers of participants in each group when performing SVC (Puechmaille 2016). The size of each subsample was based on the smallest group (rounded off to the nearest 10). A total of 30 subsamples were created, and each subsample was subjected to 1000 permutations of group membership to establish a null distribution. Permutations were performed to calculate the p-value of the prediction accuracy.

## 3 Results

Classification results are reported in Table 2 and inferential statistical comparisons are reported in Table 3. DAs are visualized in Figure 2 and CAs in Figure 3. The four best performing feature sets sorted by decoding accuracy are depicted as a confusion matrix in Figure 4.

**Figure 2.**
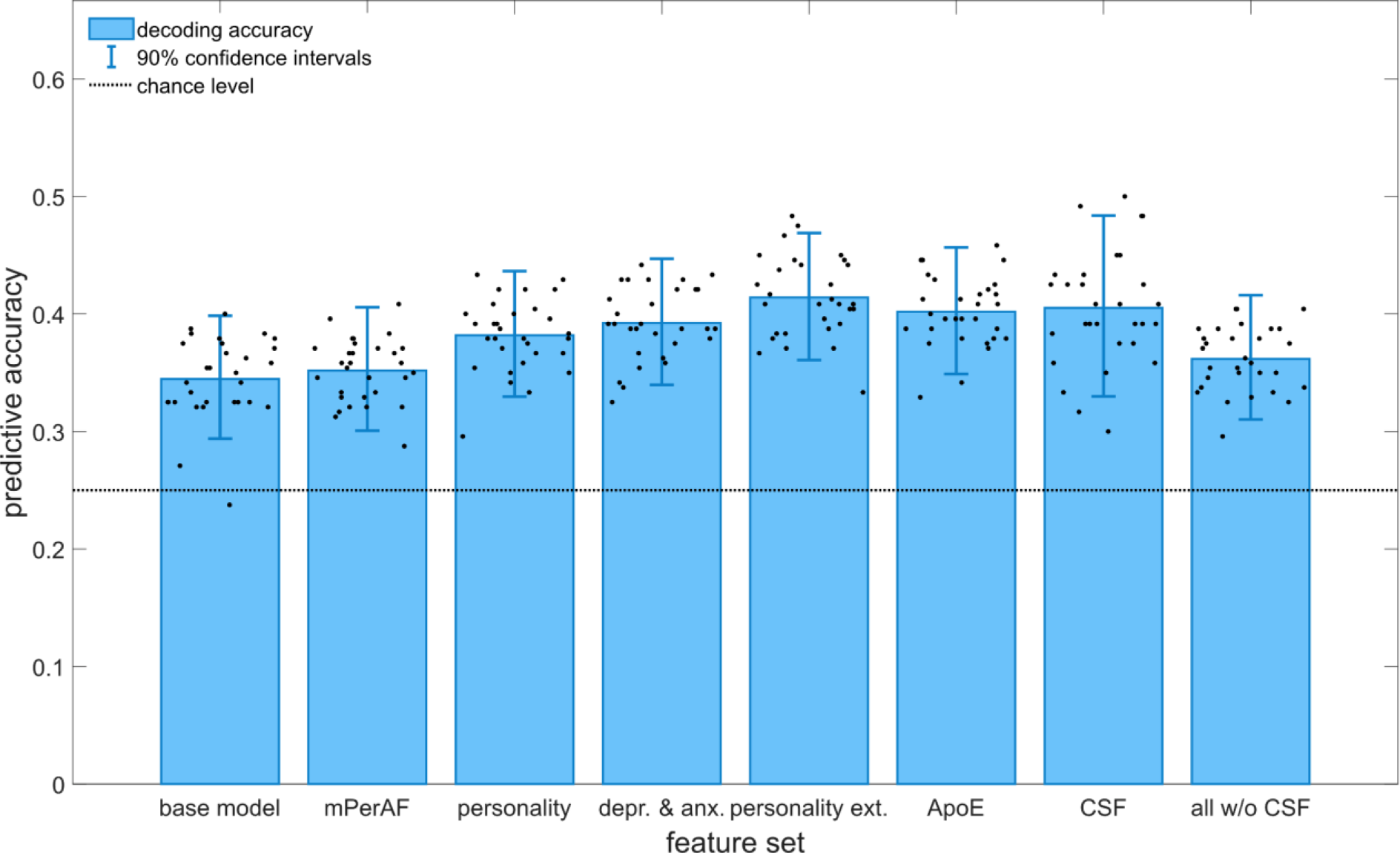
Decoding accuracies of the evaluated feature sets. The 90% confidence intervals were obtained by averaging the confidence intervals of the 30 subsamples (single dots) on which SVCs were performed.

**Figure 3.**
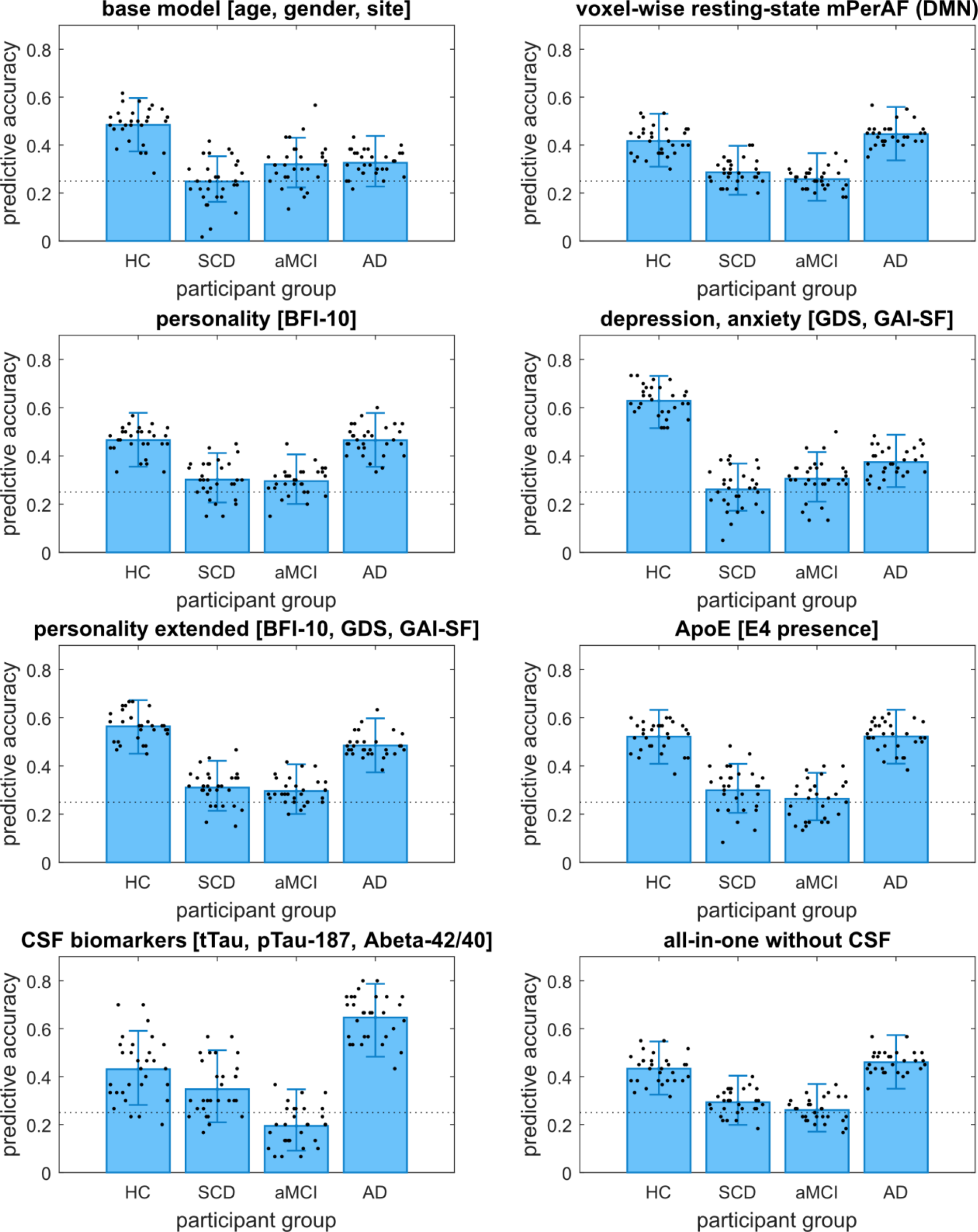
Class accuracies of the evaluated feature sets. The dotted line represents the chance level. Error bars represent the average 90% confidence interval across all 30 subsamples.

**Figure 4.**
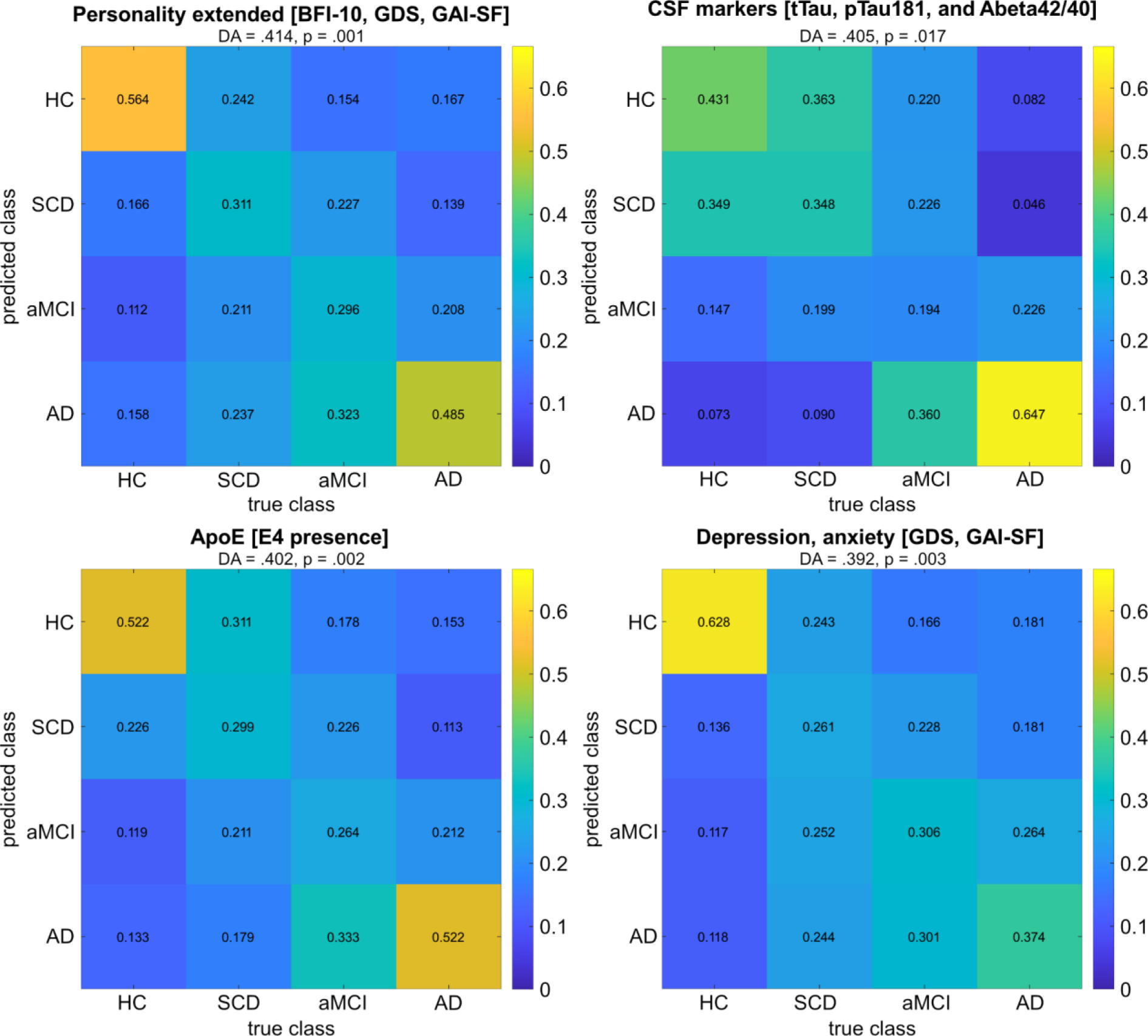
Confusion matrices of best performing feature sets by decoding accuracy.

**Table 1.** Descriptive statistics of predictor variables.

|  | HC | SCD | aMCI | AD | Statistics |
| --- | --- | --- | --- | --- | --- |
| **N** | 189 | 338 | 132 | 74 | - |
| **age range** | 60 – 87 yrs | 59 – 87 yrs | 61 – 86 yrs | 60 – 89 yrs | - |
| **mean age ± SD** | 69.09 ±5.42<br>yrs | 70.72 ±6.05<br>yrs | 72.86 ±5.61<br>yrs | 74.09 ±6.26<br>yrs | $H(3) = 52.653$,<br>$p < .001$ |
| **gender ratio** | 81/108 m/f | 183/155 m/f | 71/61 m/f | 33/41 m/f | $\chi^2(3, N = 733)$<br>$= 7.79, p = .051$ |
| **ApoE risk alleles** | N (0/1/2):<br>146/36/3 | N (0/1/2):<br>220/102/10 | N (0/1/2):<br>69/50/9 | N (0/1/2):<br>27/31/15 | $\chi^2(6, N = 718)$<br>$= 72.74, p < .001$ |
| **mean O score\*** | 3.161<br>±0.7284 | 2.888<br>±0.7872 | 3.012<br>±0.7421 | 2.948<br>±0.8263 | $H(3) = 14.249$,<br>$p = .003$ |
| **mean C score\*** | 3.196<br>±0.6841 | 3.196 ±.6645 | 3.153 ±.6539 | 2.910 ±.7067 | $H(3) = 11.917$,<br>$p = .008$ |
| **mean E score\*** | 3.175<br>±0.5097 | 3.076<br>±0.6439 | 3.129<br>±0.6744 | 3.142<br>±0.8868 | $H(3) = 4.766$,<br>$p = .190$ |
| **mean A score\*** | 3.083<br>±0.7849 | 3.148<br>±0.7328 | 3.056<br>±0.7678 | 2.758<br>±0.7708 | $H(3) = 13.769$,<br>$p = .003$ |
| **mean N score\*** | 2.825<br>±0.6475 | 2.885<br>±0.6643 | 3.077<br>±0.8166 | 3.045<br>±0.7474 | $H(3) = 10.876$,<br>$p = .012$ |
| **GDS mean / median score\*** | 0.66 / 0.00 | 2.04 / 1.00 | 2.02 / 2.00 | 2.39 / 2.00 | $H(3) = 124.69$,<br>$p < .001$ |
| **GAI-SF mean / median score\*** | 0.65 / 0.00 | 1.19 / 1.00 | 1.05 / 1.00 | 1.05 / 1.00 | $H(3) = 24.348$,<br>$p < .001$ |
| **mean tTau (pg/ml)** | 369.47<br>±148.70 | 374.20<br>±185.04 | 555.61<br>±318.78 | 791.96<br>±399.94 | $H(3) = 62.974$,<br>$p < .001$ |
| **mean pTau181 (pg/ml)** | 49.70 ±16.03 | 54.03 ±23.92 | 70.74 ±43.02 | 95.89 ±47.64 | $H(3) = 53.933$,<br>$p < .001$ |
| **mean Aβ42/40 ratio** | 0.09650<br>±0.02214 | 0.092397<br>±0.027371 | 0.073111<br>±0.030570 | 0.050423<br>±0.019247 | $H(3) = 77.923$,<br>$p < .001$ |

*Note: Demographic information along with statistics from a chi-squared test (gender ratio) and Kruskal-Wallis tests (other metrics). Abbreviations: N = sample size; f = female; m = male; O = Openness; C = Conscientiousness; E = Extraversion; A = Agreeableness; N = Neuroticism; GDS = Geriatric Depression Scale; GAI-SF = Geriatric Anxiety Index, Short Form; pg = picogram; ml = milliliter. \*see Figure S1 in the Supplementary Information*

**Table 2.** SVM classification results.

| **Feature set** | **value** | **DA** | **HC** | **SCD** | **aMCI** | **AD** |
| --- | --- | --- | --- | --- | --- | --- |
| **1. Base model** | mean accuracy | .345 | .484 | .248 | .320 | .326 |
|  | 90% CI | [.294, .398] | [.374, .596] | [.163, .353] | [.223, .431] | [.227, .438] |
|  | mean *p* | .047 | .051 | .487 | .300 | .316 |
| **2. mPerAF** | mean accuracy | .352 | .417 | .287 | .258 | .446 |
|  | 90% CI | [.301, .406] | [.310, .531] | [.193, .397] | [.168, .366] | [.336, .559] |
|  | mean *p* | .010 | .026 | .299 | .419 | .016 |
| **3. Personality** | mean accuracy | .382 | .466 | .302 | .296 | .465 |
|  | 90% CI | [.330, .436] | [.355, .579] | [.207, .412] | [.201, .406] | [.355, .578] |
|  | mean *p* | .006 | .024 | .309 | .300 | .041 |
| **4. Depression, anxiety** | mean accuracy | .392 | .628 | .261 | .306 | .374 |
|  | 90% CI | [.340, .447] | [.515, .732] | [.173, .368] | [.210, .416] | [.271, .488] |
|  | mean *p* | .003 | .003 | .448 | .311 | .186 |
| **5. Personality extended** | mean accuracy | .414 | .564 | .311 | .296 | .485 |
|  | 90% CI | [.361, .469] | [.451, .673] | [.214, .421] | [.201, .407] | [.374, .598] |
|  | mean *p* | .001 | .002 | .258 | .292 | .014 |
| **6. ApoE** | mean accuracy | .402 | .522 | .299 | .264 | .522 |
|  | 90% CI | [.349, .457] | [.409, .633] | [.206, .409] | [.175, .372] | [.410, .633] |
|  | mean *p* | .002 | .021 | .342 | .445 | .023 |
| **7. CSF** | mean accuracy | .405 | .431 | .348 | .194 | .647 |
|  | 90% CI | [.330, .484] | [.282, .591] | [.210, .510] | [.091, .347] | [.483, .787] |
|  | mean *p* | .017 | .156 | .301 | .675 | .009 |
| **8. All w/o CSF** | mean accuracy | .362 | .433 | .293 | .261 | .460 |
|  | 90% CI | [.310, .416] | [.325, .547] | [.199, .404] | [.170, .369] | [.350, .573] |
|  | mean *p* | .006 | .016 | .265 | .416 | .012 |

*Note.* Because four groups were included, chance performance was at .25. Mean accuracy, 90% CI and mean *p* correspond to the average across 30 subsamples. The *p*-value of each subsample was obtained by comparing the accuracy value to the null distribution generated from 1000 permutations.

**Table 3.**
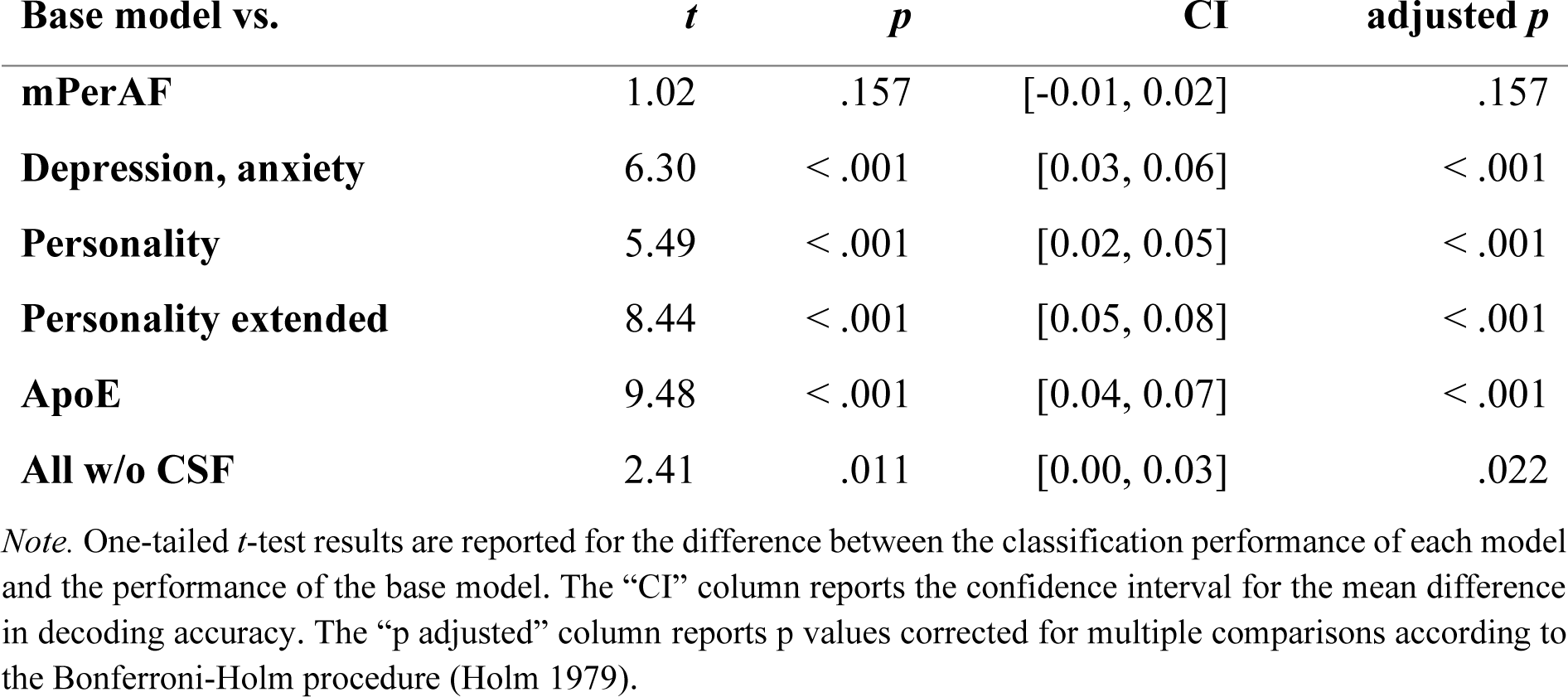
Inferential statistical comparisons of decoding accuracy between feature sets.

### 3.1 Base model: Low predictive value of combining age, gender, and site

The “base model” produced the lowest overall DA (DA = .345, *p* = .047) and no CA was significantly different from chance for any group (Figure 3).

### 3.2 mPerAF: Low but above-chance performance of resting-state DMN activity

Feature set “mPerAF” performed significantly above chance (DA = .352, p = .010), along with significant CAs for both HC (CA = .417, p = .026) and AD (CA = .446, p = .016). CAs for SCD (CA = .287, *p* = .299) and aMCI (CA = .258, *p* = .419) were statistically indifferent from chance.

### 3.3 Personality trait and affective state scores: Highest prediction accuracies for HC and across groups

Feature set “Personality” was consistently outperformed by “Personality extended”, which produced the overall highest DA (DA = .414, *p* = .001). Combining scores of geriatric depression and anxiety demonstrated the overall highest class accuracy for healthy controls (CA = .628, *p* = .003) and the overall third-highest DA (.392, *p* = .003).

### 3.4 ApoE: Third-highest decoding accuracy

Feature set “ApoE” showed the third-best performance (DA = .402, *p* = .002). It also demonstrated significantly above chance CAs for HC (CA = .522, *p* = .021) and AD (CA = .522, *p* = .023).

### 3.5 Relatively poor performance of combined predictors without CSF biomarkers

Across all groups and in terms of DA, prediction accuracies of feature set “All w/o CSF” were consistently lower than those of “Personality” and “Personality extended” and it was not in the top three CAs for any participant group.

### 3.6 CSF biomarkers predict AD best, but perform poorly for HC

Feature set “CSF” exhibited the highest CAs for the groups of SCD (CA = .348, *p* = .301) and AD (CA = .647, *p* = .009), as well as the second-highest DA (.405, *p* = .017). CAs for HC (CA= .431, *p* = .156) and aMCI (CA = .194, *p* = .675) were non-significant above chance.

### 3.7 Comparison of feature sets and summary

The highest performance in terms of DA (Figure 4) were achieved by the feature sets “Personality extended” (DA = .414, *p* = .001) followed by “CSF” (DA = .405, *p* = .017), “ApoE” (DA = .402, *p* = .002), and “Depression, anxiety” (DA = .392, *p* = .003). All feature sets—except “mPerAF”—performed significantly better than the base model in predicting group membership (Table 3).

## 4 Discussion

In this cross-sectional study, we aimed to evaluate the diagnostic value of several feature sets for Alzheimer’s disease, associated at-risk states (SCD, aMCI), and healthy controls using support vector machine classification. We focused on the performance of combining personality traits with scores of depression and anxiety, as well as examining the predictive ability of DMN BOLD amplitude fluctuation measured through resting-state fMRI, ApoE genotype, and CSF biomarkers. All feature sets demonstrated decoding accuracy significantly above chance (Table 2).

The highest decoding accuracy was observed in feature sets: (i) “Personality extended,” which combined personality traits with anxiety and depression scores; (ii) “CSF”, consisting of tTau, pTau181, and Aβ42/40 ratio; (iii) “ApoE,” including the ApoE genotype; and (iv) “Depression, anxiety,” comprising depression and anxiety scores. The only feature sets not achieving significant above-chance classification performance for HC were “Base model” and “CSF”, with the latter showing the lowest overall accuracy for the aMCI group.

### 4.1 Inferiority of the combined predictor and poor prediction accuracy of resting-state activity of the DMN

Our hypothesis that combining non-invasive predictors (feature set “All w/o CSF”) would outperform CSF biomarkers in prediction accuracy was not supported by our data. The classification accuracies of the “All w/o CSF” feature set were comparably low and similar to the “mPerAF” feature set, suggesting that the inclusion of mPerAF paradoxically reduced classification performance. While DMN resting-state mPerAF performed above chance, its performance did not significantly differ from the “Base model”.

The predictive ability of resting-state fMRI of the DMN for AD has yielded inconsistent findings. While certain studies have reported consistent alterations in DMN activity and connectivity in AD (Mevel et al. 2011) and the added value of combining different MRI modalities to classify AD (Schouten et al. 2016), other research suggests that neuropsychiatric measures may have higher predictive ability (Gill et al. 2020).

It is important to note that most DMN studies have focused on functional connectivity rather than voxel-wise amplitude measures like mPerAF. The divergent results could be attributed to our approach of evaluating all groups simultaneously, resembling a fully automated diagnostic process, as opposed to making binary decisions between distinct groups. Furthermore, unequal sample sizes can introduce bias in classification, and various approaches have been proposed to address this issue (Jo et al. 2019).

### 4.2 A combination of personality, anxiety, and depression scores yield a relatively high overall prediction accuracy

Personality alone demonstrated class accuracies statistically significant above chance for the groups of HC and AD, but not for SCD and aMCI, partially confirming our hypothesis. “Personality” was surpassed by the feature set “Personality extended”. However, the accuracy of correctly classifying the aMCI group was equally high, while class accuracies for the SCD and aMCI groups remained nonsignificant, partially supporting our hypothesis. These results indicate that depression and anxiety contribute additional predictive value to the decoding accuracy of the BFI-10. The highest class accuracy for HC, however, was achieved by a feature set containing scores of depression and anxiety, and adding personality traits did not improve class accuracy. Previous studies have indicated that depressive episodes can be prodromal manifestations of neurodegeneration in AD (Panza et al. 2010; Singh-Manoux et al. 2017; Hansen et al. 2021). Possibly, alterations in levels of depression within the SCD and aMCI groups surpass changes in personality traits when contrasted with shifts seen in healthy controls. The predictive ability of the feature set “Depression, anxiety” for HC may be primarily attributed to the GDS as some of the GAI-SF items overlap with those of the BFI-10 neuroticism scale, suggesting depression scores to be well-suited in distinguishing between healthy individuals and participants with cognitive impairment. AD participants were best classified using a combination of CSF biomarkers, consistent with previous findings (Olsson et al. 2016; Lleó et al. 2019; Düzel et al. 2022). The predictive value of combining CSF biomarkers, personality traits and scores of depression and anxiety should be investigated further.

### 4.3 Poor classification accuracy for SCD and aMCI with any feature set

Predictions for participant groups with SCD or aMCI were mostly above chance level but not statistically significant (Table 2). This trend persisted after merging SCD and aMCI into an “at-risk for AD” group (Table S2). Neither SCD nor aMCI are specific to AD and can be caused by a variety of conditions, including normal aging. Because the underlying conditions causing SCD or aMCI in DELCODE participants were not assessed at the study’s outset, it is reasonable to assume that a proportion of participants did not actually have preclinical AD (see section 2.1). Identification of those individuals with SCD or aMCI not due to AD likely largely failed as we used predictors that are specific to AD, explaining the poor class accuracies for the groups of SCD, aMCI, and “at-risk for AD” (Chételat et al. 2005; Johns et al. 2012; Bessi et al. 2018).

### 4.4 Limitations

Our study has several limitations. CSF biomarkers were only measured in a portion of the sample, resulting in different sample sizes for feature sets and exclusion of the “CSF” feature set from inferential analysis. Anosognosia is known to be a common occurrence in the early stages of AD (Leicht et al. 2010; Orfei et al. 2010; de Ruijter et al. 2020) and may also have confounded the assessments of the GDS, the GAI-SF (Starkstein 2014), and the BFI-10 (Agϋera-Ortiz et al. 2019). Additionally, caregiver influence on self-reports may have affected the accuracy of assessments in the aMCI and AD groups. Another important limitation relates to the demographics of the groups. Despite being composed of confounding variables only, the “Base model” performed above chance. This can be attributed to the association between age and dementia risk (Terracciano & Sutin 2019). On average, AD participants were older than HC or those with SCD (Table 1). However, because age was included in all feature sets, its predictive value was consistently accounted for. Finally, the cross-sectional design is a limitation, as it precludes to track personality change across time and assess the validity of the markers over the natural progression of the participants. This underscores the need for future research to complement our findings with longitudinal data.

### 4.5 Conclusions

Our results show that no single combination of the evaluated features achieved consistently superior class accuracies for all assessed participant groups. The combination of depression and anxiety scores was most effective in classifying healthy controls, supporting previous findings that regard late-life depression as a prodrome of Alzheimer’s disease, while CSF biomarkers were most effective in classifying participants with mild Alzheimer’s disease. The highest overall prediction accuracies across all participant groups were achieved by a combination of personality traits with scores of depression and anxiety, closely followed by CSF biomarkers and the ApoE genotype. These findings indicate that a combination of CSF biomarkers, personality, depression and anxiety scores, and the ApoE genotype may have complementary value for classification of AD and associated at-risk states. Further investigation is needed, particularly regarding the predictive value of personality traits and associated affective states as low-cost and easily assessable screening tools. Moreover, our findings highlight the challenge of accurately classifying SCD and aMCI groups using machine learning approaches when the underlying conditions of these cognitive impairments are unknown. Addressing this challenge requires adhering to consensus on terminology and conceptual frameworks.

## Data availability statement

All scripts (https://github.com/jmkizilirmak/DELCODE162) and the machine learning toolbox for Matlab (https://github.com/JoramSoch/ML4ML) are available online. Data, study protocol, and biomaterials can be shared with collaborators based on individual data and biomaterial transfer agreements with the DZNE.

## Ethics approval statement

The study protocol was approved by the Institutional Review Boards of all participating study centers of the DZNE. The process was led and coordinated by the ethical committee of the medical faculty of the University of Bonn (registration number 117/13).

## Patient consent statement

All participants gave written informed consent.

## Permission to reproduce material from other sources

Not applicable.

## Clinical trial registration

The DELCODE study has been registered as a clinical trial with the German Clinical Trials Register under the study acronym “DELCODE”, ID DRKS00007966.

## Supporting information

Supplementary Results

## Data Availability

All scripts used to perform the analyses are available under https://github.com/jmkizilirmak/DELCODE162. Data can be made available to cooperation partners of the DZNE after setting up appropriate data sharing contracts.

## 5 List of Abbreviations

Aβ: Amyloid beta
AD: Alzheimer’s disease
aMCI: amnestic mild cognitive impairment
ANOVA: analysis of variance
BFI: Big Five Inventory
BFI-10: Big Five Inventory 10-item short form
BOLD: blood oxygenation level-dependent
CERAD: Consortium to Establish a Registry for Alzheimer’s Disease
CA: class accuracy
CI: confidence interval
CSF: cerebrospinal fluid
DA: decoding accuracy
DMN: default mode network
DELCODE: DZNE-Longitudinal Cognitive Impairment and Dementia Study
DZNE: Deutsches Zentrum für Neurodegenerative Erkrankungen (English: German Center for Neurodegenerative Diseases)
EPI: echo-planar imaging
fMRI: functional magnetic resonance imaging
FWHM: full width at half maximum
GAI-SF: Geriatric Anxiety Inventory, Short Form
GDS: Geriatric Depression Scale
HC: healthy controls
Hz: Hertz
MCI: mild cognitive impairment
NIA: National Institute on Aging
MMSE: Mini Mental Status Examination
MNI: Montreal Neurological Institute
mPerAF: mean percent amplitude of fluctuation
MPRAGE: Magnetization Prepared Rapid Gradient Echo
MRI: magnetic resonance imaging
NEO PI-R: Revised NEO Personality Inventory
PerAF: percent amplitude of fluctuation
pTau181: phosphorylated tau181
ROI: region of interest
rs-fMRI: resting-state functional magnetic resonance imaging
SCD: subjective cognitive decline
SD: standard deviation
SPM: Statistical Parametric Mapping
SVC: support vector classification
SVM: support vector machine
TE: echo time
TR: time to repetition
tTau: total tau
VDM: voxel-displacement map
yrs: years

## 6 Acknowledgments

We would like to thank all the participants in the DELCODE study and all the technical, medical, and psychological staff without whom this study would not have been possible. Special thanks go to the MRI centers at the Max-Delbrück-Center for Molecular Medicine (MDC) of the Helmholtz Association, the Center for Cognitive Neuroscience Berlin (CCNB) at the Free University of Berlin, and the Bernstein Center for Computational Neuroscience (BCCN), Berlin.

## Footnotes

1 In this study, the term “predictive” refers to support vector classification performance of feature sets differentiating participant groups in a cross-sectional design, not the prediction of a longitudinal diagnostic outcome.

2 Since the *RESTplus* toolbox only provides four default masks, a group-level mask fitting the dimensions and voxel sizes of our pre-processed task-based fMRI was generated and added to the mask directory. Additionally, the parallel processing mode using outdated MATLAB commands had to be turned off.

