## Supplementary Results for "Machine learning-based classification of Alzheimer’s disease and its at-risk states using personality traits, anxiety, and depression"

#### Abbreviations

| | |
|---|---|
| AD | Alzheimer's disease |
| BFI | Big Five inventory |
| CA | class accuracy |
| CSF | cerebrospinal fluid |
| DA | decoding accuracy |
| HC | healthy controls |
| aMCI | amnestic mild cognitive impairment |
| SCD | subjective cognitive decline |
| SVC | support vector classification |
| SVM | support vector machine |

### 1. Supplementary Methods

#### 1.1. Participants

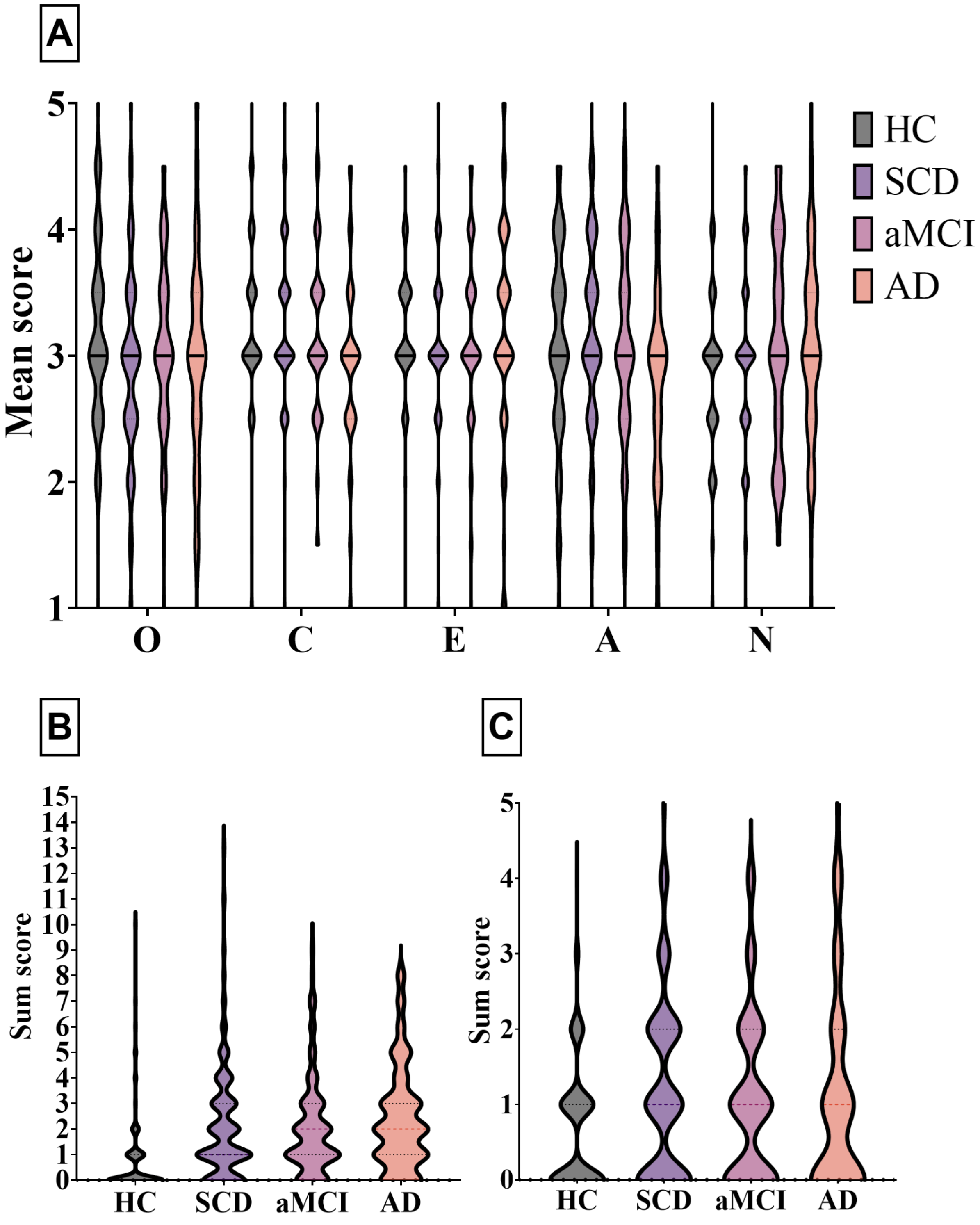


**Figure S1. Violin plots of psychometric scores.** A. Mean BFI-10 scores (O = Openness; C = Conscientiousness; E = Extraversion; A = Agreeableness; N = Neuroticism). B. GDS sum scores. C. GAI-SF sum scores.

#### 1.2. Predictor variables

**Table S1. Overview of predictor variables**

| Predictor variable(s) | Range | Description |
|---|---|---|
| Age | $59 \le x_i \le 89$ | chronological age in years |
| Gender | $x_i \in \{0,1\}$ | male and female were one-hot-encoded as separate variables |
| Site | $x_{ij} \in \{0,1\}$ | each of 10 sites was one-hot-encoded as a separate variable |
| Resting-state DMN activity | $x_i \in \mathbb{R}^v$ | mPerAF maps masked for DMN |
| BFI:<br>– Neuroticism<br>– Extraversion<br>– Openness<br>– Conscientiousness<br>– Agreeableness | $1 \le x_i \le 5$ | each trait was assessed as the mean of the rating on two items (5-point scales) of the 10-item BFI |
| Depression | $0 \le x_i \le 13$ | GDS sum score |
| Anxiety | $0 \le x_i \le 5$ | GAI-SF sum score |
| ApoE genotype | $x_i \in \{0, 1, 2\}$ | number of ε4 (risk) alleles |
| CSF: | | |
| – tTau | $66.4 \le x_i \le 2067.5$ | unit: pg/ml |
| – pTau181 | $14.92 \le x_i \le 320.62$ | unit: pg/ml |
| – Aβ42/40 ratio | $0.026 \le x_i \le 0.151$ | - |

#### 1.3. Supplementary Results

#### 1.4. Support vector classification with SCD and aMCI as one single "risk group"

In the main paper, we reported that every feature set yielded low CAs for the at-risk states SCD and aMCI. Therefore, we decided to combine the groups of SCD and aMCI participants into a common group of "at-risk for AD" and repeated SVM classifications for all feature sets as described in the main paper. Sample sizes were: 179 HC, 421 at-risk for AD, 63 AD.

**Table S2. SVM classification results**

| Feature set | Value | DA | HC | at-risk for AD | AD |
|---|---|---|---|---|---|
| **1. Base model** | mean accuracy | .444 | .532 | .410 | .389 |
| | 90% CI | [.381, .508] | [.419, .642] | [.305, .522] | [.284, .503] |
| | mean *p* | .045 | .081 | .298 | .361 |
| **2. mPerAF** | mean accuracy | .457 | .461 | .358 | .552 |
| | 90% CI | [.394, .520] | [.351, .573] | [.256, .471] | [.438, .661] |
| | mean *p* | .033 | .132 | .365 | .005 |
| **3. Personality** | mean accuracy | .481 | .496 | .406 | .541 |
| | 90% CI | [.418, .545] | [.384, .607] | [.300, .519] | [.428, .651] |
| | mean *p* | .007 | .088 | .254 | .031 |
| **4. Depression, anxiety** | mean accuracy | .501 | .653 | .399 | .450 |
| | 90% CI | [.437, .564] | [.540, .754] | [.294, .512] | [.341, .563] |
| | mean *p* | .003 | .007 | .304 | .188 |
| **5. Personality extended** | mean accuracy | .527 | .611 | .410 | .559 |
| | 90% CI | [.463, .590] | [.498, .716] | [.304, .523] | [.446, .668] |
| | mean *p* | .001 | .004 | .222 | .020 |
| **6. ApoE** | mean accuracy | .532 | .577 | .389 | .631 |
| | 90% CI | [.468, .59] | [.464, .68] | [.285, .50] | [.518, .73] |
| | mean *p* | .001 | .043 | .328 | .010 |
| **7. CSF** | mean accuracy | .551 | .521 | .367 | .767 |
| | 90% CI | [.460, .640] | [.361, .678] | [.224, .530] | [.607, .883] |
| | mean *p* | .006 | .119 | .433 | .002 |
| **8. All w/o CSF** | mean accuracy | .466 | .472 | .368 | .558 |
| | 90% CI | [.403, .530] | [.362, .584] | [.266, .481] | [.444, .667] |
| | mean *p* | .028 | .112 | .318 | .004 |

*Note.* Since three groups were included, the chance level was at 33.33%. Mean accuracy and mean *p* refer to the mean of 30 subsamples. The *p*-value of each subsample was obtained by comparing the accuracy value to the null distribution generated from 1000 permutations.

**Table S3. Inferential statistical comparisons of DA between feature sets**

| Base model vs. | t | p | CI | p adjusted |
|---|---|---|---|---|
| **mPerAF** | 1.16 | .128 | [-0.01, 0.03] | .128 |
| **Depression, anxiety** | 7.37 | < .001 | [0.04, 0.07] | < .001 |
| **Personality** | 6.85 | < .001 | [0.03, 0.05] | < .001 |
| **Personality extended** | 9.94 | < .001 | [0.07, 0.10] | < .001 |
| **ApoE** | 9.85 | < .001 | [0.07, 0.10] | < .001 |
| **All w/o CSF** | 1.96 | .030 | [0.00, 0.04] | .060 |

*Note. One-tailed t-test results are reported for the difference between the classification performance of each model and the performance of the base model. The column "p adjusted" reports p-values corrected for multiple comparisons according to the Bonferroni-Holm procedure (Holm, 1979).*

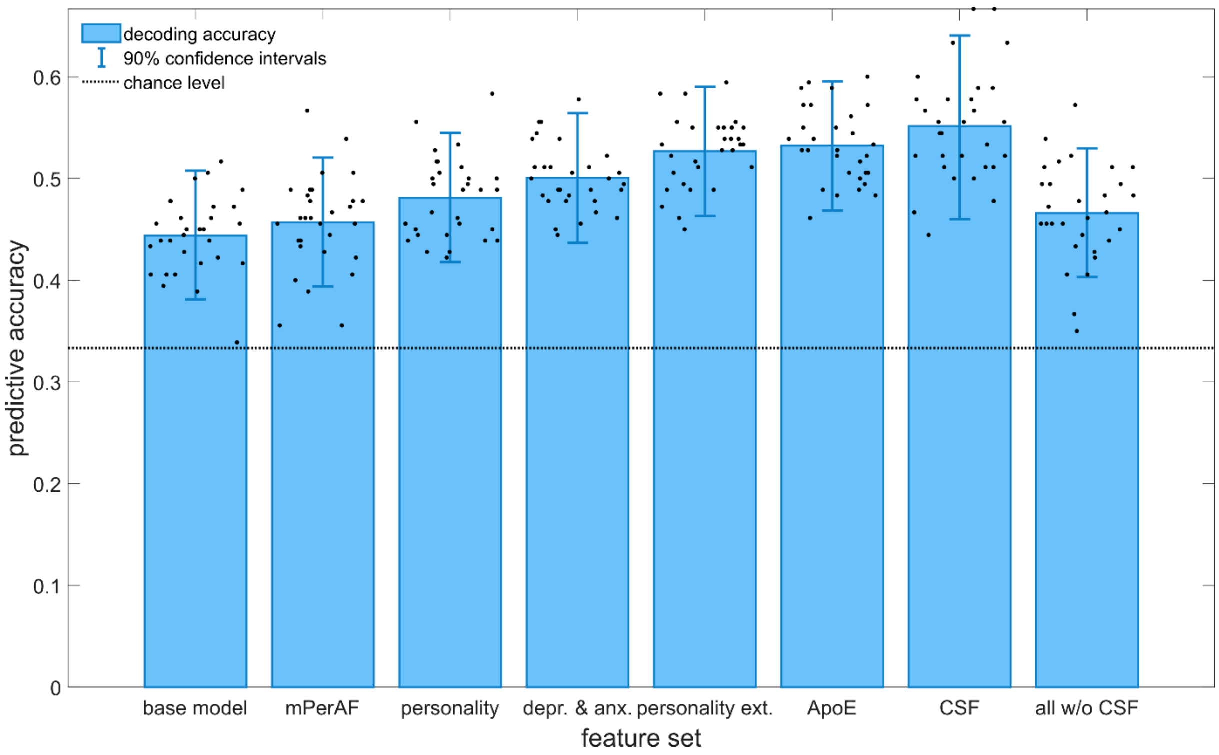


**Figure S2. Decoding accuracies for the evaluated feature sets.** Here, SCD and aMCI were combined into an "at-risk for AD" group. Error bars represent 90% confidence intervals obtained by averaging the confidence intervals of the 30 subsamples (single dots) on which SVCs were performed.

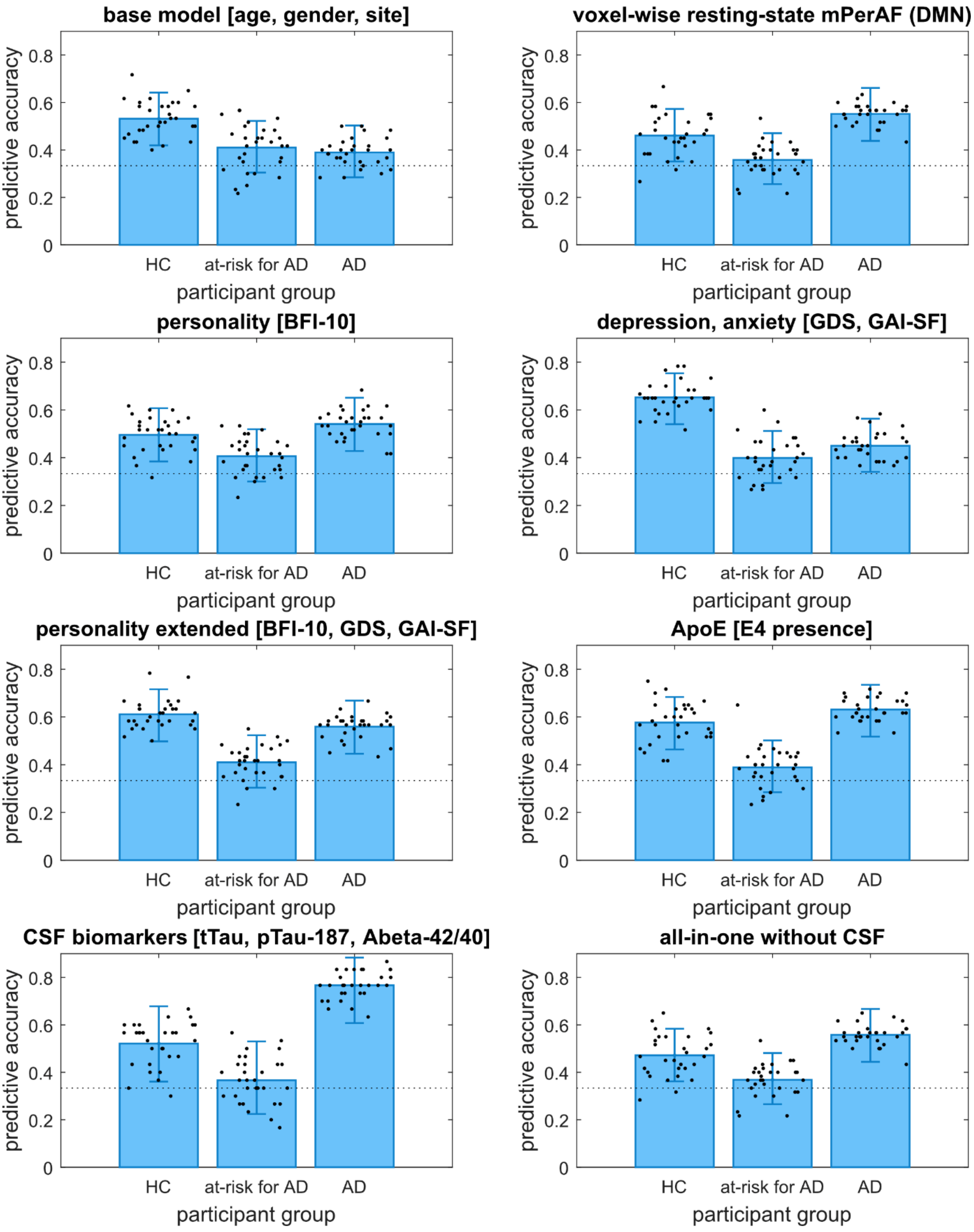


**Figure S3. Class accuracies of the evaluated feature sets.** The “at-risk for AD” group is composed of SCD and aMCI participants. The dotted line represents the chance level, and the error bars represent the average 90% confidence interval across all 30 subsamples.

##### 1.5. Support vector classification with the same sample size for all feature sets

To allow an inferential statistical comparison of the decoding accuracies of all feature sets – including "CSF" – we ran another set of SVCs with a sample that included only participants who had no missing data in any of the features. This resulted in a reduced but equal sample size of 311 participants (74 HC, 142 SCD, 63 aMCI, 32 AD) across all feature sets. This time, feature set number eight consisted of all predictors, including CSF biomarkers. The results are shown in Table S4.

**Table S4. SVM classification results for the reduced sample (all feature sets with N = 311)**

| Feature set | value | DA | HC | SCD | aMCI | AD |
|---|---|---|---|---|---|---|
| **1. Base model** | mean accuracy | .350 | .339 | .320 | .249 | .492 |
| | 90% CI | [.278, .428] | [.201, .502] | [.188, .480] | [.131, .407] | [.333, .653] |
| | mean *p* | .064 | .302 | .373 | .511 | .073 |
| **2. mPerAF** | mean accuracy | .349 | .332 | .248 | .293 | .522 |
| | 90% CI | [.277, .426] | [.197, .494] | [.127, .408] | [.164, .455] | [.361, .680] |
| | mean *p* | .095 | .300 | .550 | .410 | .015 |
| **3. Personality** | mean accuracy | .363 | .387 | .364 | .229 | .472 |
| | 90% CI | [.290, .441] | [.241, .551] | [.223, .527] | [.115, .386] | [.315, .634] |
| | mean *p* | .041 | .156 | .230 | .578 | .065 |
| **4. Depression, anxiety** | mean accuracy | .377 | .546 | .312 | .217 | .434 |
| | 90% CI | [.303, .456] | [.383, .701] | [.181, .473] | [.108, .371] | [.281, .598] |
| | mean *p* | .020 | .026 | .384 | .613 | .128 |
| **5. Personality extended** | mean accuracy | .384 | .489 | .352 | .237 | .460 |
| | 90% CI | [.310, .463] | [.331, .649] | [.212, .515] | [.121, .395] | [.304, .622] |

| | | | | | | |
|---|---|---|---|---|---|---|
| | mean *p* | .024 | .040 | .231 | .558 | .097 |
| **6. ApoE** | mean accuracy | .381 | .457 | .284 | .184 | .598 |
| | 90% CI | [.307, .459] | [.302, .618] | [.160, .444] | [.086, .335] | [.433, .747] |
| | mean *p* | .027 | .108 | .459 | .685 | .016 |
| **7. CSF** | mean accuracy | .419 | .448 | .368 | .193 | .669 |
| | 90% CI | [.344, .498] | [.295, .609] | [.228, .528] | [.090, .347] | [.504, .807] |
| | mean *p* | .019 | .100 | .263 | .684 | .002 |
| **8. All-in-one (incl. CSF)** | mean accuracy | .361 | .358 | .259 | .289 | .539 |
| | 90% CI | [.289, .439] | [.218, .520] | [.136, .420] | [.160, .451] | [.376, .695] |
| | mean *p* | .063 | .239 | .504 | .414 | .010 |

*Note.* Since four groups were included, the chance performance was at 25%. Mean accuracy, mean *p* refers to the mean across 30 subsamples. The *p*-value of each subsample was obtained by comparing the accuracy value to the null distribution generated from 1000 permutations.

**Table S5. Inferential statistical comparisons of DA between feature sets**

| **CSF only vs.** | **t** | **p** | **CI** | **p adjusted** |
|---|---|---|---|---|
| **Base model** | 7.76 | < .001 | [0.05, 0.08] | < .001 |
| **mPerAF** | 7.67 | < .001 | [0.05, 0.09] | < .001 |
| **Depression, anxiety** | 4.34 | < .001 | [0.02, 0.06] | < .001 |
| **Personality** | 5.38 | < .001 | [0.04, 0.08] | < .001 |
| **Personality extended** | 3.00 | .003 | [0.01, 0.06] | .003 |
| **ApoE** | 4.19 | < .001 | [0.02, 0.05] | < .001 |
| **All-in-one** | 6.52 | < .001 | [0.04, 0.07] | < .001 |

Compared to the larger sample size variant, the overall performance ranking of the feature sets was slightly different (see main paper), as can be seen in Table S4 and Figure S4. Feature set “CSF” yielded the overall highest decoding accuracy (DA = .419, $p$ = .019). Inferential statistical comparisons are reported in Table S5.

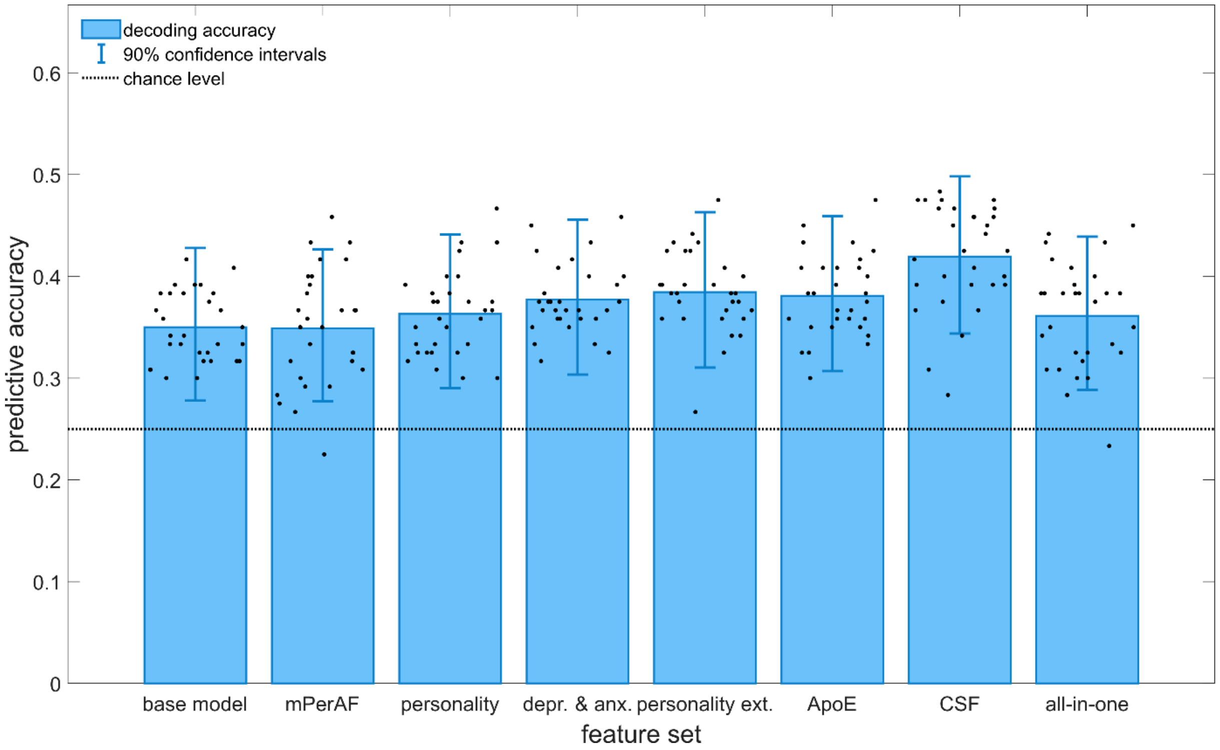


**Figure S4. Decoding accuracies for the evaluated feature sets with reduced but equal sample size (N = 311).**

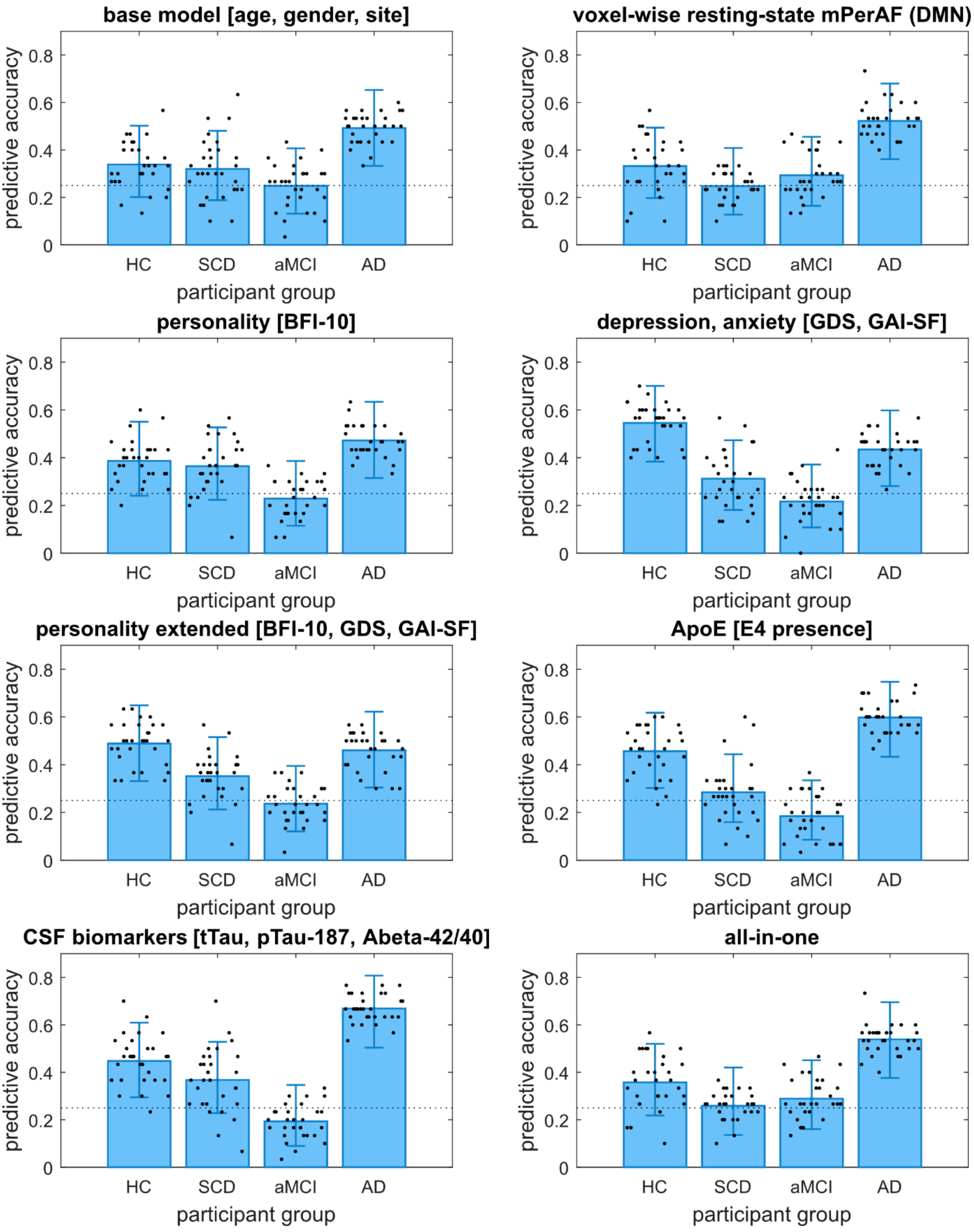


**Figure S5. Class accuracies for the evaluated feature sets with reduced but equal sample size for all (N = 311)**.

#### 2. Supplementary Discussion

##### 2.1. Combining SCD and aMCI into an "at-risk for AD" group does not meaningfully improve class accuracies

We found no meaningful change in the performance pattern of the best performing feature sets (Table S2, Figure S2). The three best performing feature sets in terms of mean DA were "CSF" (DA = .551, *p* = .006), "ApoE" (DA = .532, *p* = .001), and "Personality extended" (DA = .527, *p* = 001). The CAs for the "at-risk for AD" group remained poor and statistically non-significant across all feature sets. We again observed that feature set "Depression, anxiety" had the highest CA for healthy participants (CA = .653, *p* = .007), while feature set "CSF" performed best in classifying AD patients correctly (CA = .767, *p* = .002). Notably, the only two feature sets achieving CAs statistically significantly above chance level for both groups of HC and AD were "Personality extended" and "ApoE" (Table S2, Figure S3).

According to one-tailed pairwise comparisons between each feature set and the "Base model" (see Table S3), all feature sets except for "mPerAF" and "All w/o CSF" performed statistically significantly better than the "Base model".

##### 2.2. Equal sample sizes across all feature sets: CSF biomarkers achieve the highest overall decoding accuracy and class accuracy for AD

In the main paper, we reported that feature sets "Personality extended" and "CSF" had almost equal decoding accuracies. In the variant with equal sample sizes, the DA of "CSF" increased relative to other feature sets and achieved the overall highest decoding accuracy (DA = .419, *p* = .019). However, while "CSF" also yielded the highest class accuracy for the AD group (CA = .669, *p* = .002), this is contrasted by statistically non-significant CAs for HC, SCD, and aMCI. Feature set "Depression, anxiety" again achieved the highest CA for HC (CA = .546, *p* = .026).

Prediction results from the smaller-but-equal sample size variant reemphasized that CSF biomarkers, psychometric scores, and the ApoE genotype likely have complementary value, as they provide decent prediction accuracies for different participant groups.
